# Thalamocortical seizure onset patterns in drug resistant focal epilepsy

**DOI:** 10.1101/2025.04.11.25325282

**Authors:** Hugh D. Simpson, Vaclav Kremen, Vladimir Sladky, Benjamin H. Brinkmann, Nicholas M. Gregg, Brian N. Lundstrom, Kai J. Miller, Jamie J. Van Gompel, Greg A. Worrell

## Abstract

Drug-resistant epilepsy affects tens of millions of people worldwide and is associated with considerable morbidity and mortality. Thalamic deep brain stimulation and cortical responsive neurostimulation are proven treatments for focal epilepsy. Both have been used to target a range of thalamic nuclei, yet the roles of these thalamic nuclei in focal seizure generation remain incompletely understood.

Thirteen patients with drug-resistant focal epilepsy undergoing intracranial EEG were consented to undergo investigation of thalamocortical networks. Sampled regions included cortical, mesial temporal, and thalamic brain regions. Visual and spectral analyses were performed to identify seizure onset patterns and correlate thalamic and cortical seizure activity.

Thalamic ictal discharges were observed in all patients, including synchronous thalamocortical seizure onset discharges with distinct onset patterns. These onset patterns ranged from hypersynchronous spiking, low-voltage fast activity, ictal baseline shifts, to broadband suppression. Multiple thalamic nuclei were involved in ictal organization and propagation, with the specific nuclei depending on the cortical seizure network.

The thalamus plays a crucial role in focal onset seizure generation and propagation, with distinct seizure onset patterns and nuclei involved. These findings support exploring a broader range of thalamic nuclei in epilepsy neurostimulation and have implications for seizure detection settings in intracranial sensing devices.

## Introduction

Drug-resistant epilepsy (DRE) affects up to one-third of individuals with epilepsy worldwide and is associated with substantial morbidity and mortality.^1^ While resective or ablative surgery remains the most effective treatment for appropriately selected patients with focal epilepsy, many are poor candidates for such interventions, while some who undergo surgery may yet continue to experience seizures. For these individuals, electrical brain stimulation (EBS) offers a promising adjunctive therapy.

EBS modalities, including deep brain stimulation (DBS), responsive neurostimulation (RNS), and vagus nerve stimulation (VNS), are effective in reducing seizure frequency, with median reductions of up to 75%.^2–4^ Additionally, some patients achieve seizure freedom for months to years. Among these techniques, DBS of the anterior nucleus of the thalamus (ANT) has Class I evidence for efficacy in focal epilepsy.^2,3^ Other thalamic nuclei, such as the centromedian nucleus (CMT), show promise for generalized and multifocal epilepsy.^5–7^ Emerging evidence also highlights the potential of pulvinar nucleus (PUL) DBS in focal epilepsy, though its mechanisms remain underexplored.^8–10^ Despite these advances, significant gaps persist in our understanding of thalamocortical networks and their role in seizure generation (ictogenesis) and propagation. Moreover, recent studies challenge traditional assumptions about thalamocortical circuitry, such as the surprising efficacy of ANT-DBS in generalized epilepsy,^11^ and the use of CMT-DBS in focal epilepsy.^12,13^ Consequently, many facets of EBS, including optimal patient selection, target choice for specific epilepsy syndromes, and stimulation parameters, remain poorly defined.

The role of the thalamus in ictogenesis and epileptogenesis is a topic of continued interest.^9,14–16^ Intracranial local field potential (LFP) recordings suggest that various thalamic nuclei—including the ANT, mediodorsal thalamus (DMT), and PUL—are recruited during seizure onset and propagation.^17,18^ This is of particular relevance for programming accurate seizure detectors using sensing-capable DBS devices^19^ and RNS systems. While originally used exclusively for cortical seizure detection and responsive stimulation in hippocampus and neocortex, there is growing interest in targeting thalamic nodes with RNS.^8^ Thus the timing and patterns of thalamic involvement in ictogenesis become crucial for programming detectors. Often, seizure detection is cortically based, while stimulation may be cortical and/or thalamic. This is based on the assumption that cortex leads thalamus in seizure onset in focal epilepsy,^20^ as opposed to the simultaneous thalamocortical dynamics seen in some generalised epilepsies.^21^ The possibility of synchronous thalamocortical networks in focal epilepsy would challenge this dogma and approach. Finally, while cortical seizure onset patterns are well-characterized,^15,22^ thalamocortical network correlates of these patterns is only beginning to be understood.^23^

In this study, we aim to advance our understanding of the thalamus’s role in ictogenesis by analyzing thalamocortical network dynamics during focal seizures.

## Materials and methods

### Study Population

Thalamocortical seizure onsets were studied in 13 patients with drug-resistant focal epilepsy. Two groups were included. The first was made up of 10 patients undergoing stereo-electroencephalography (sEEG) as part of routine pre-surgical evaluation, who had a single electrode trajectory extended to sample the thalamus, where it was safe and feasible to do so. This was performed under Institutional Review Board (IRB) ethical approval (IRB 15-007984). The second group was made up of three patients implanted with an investigational neural sensing and stimulation device (RC+S^TM^) with bilateral ANT and mesial temporal leads; under FDA Investigational Device Exemption (G180224) and IRB approval (IRB 18-005483). All participants provided informed consent. Clinical characteristics and outcomes were reviewed retrospectively.

### Data analysis

Seizure onset and propagation patterns were analyzed visually and quantitatively using custom MATLAB scripts. In the RC+S^TM^ group, automatic seizure detection was used to identify candidate seizures first, as described previously.^24^ In both groups seizures were confirmed using visual review by trained epileptologists. Thalamocortical ictal circuits were characterized based on the timing and spectral patterns of thalamic discharges relative to cortical SOZ activity.

Preoperative 3T MRI was co-registered with postoperative CT scans for localization of electrode contacts. Lead DBS was used to identify thalamic nuclei.^25^ Groupings of thalamic nuclei included: anterior (ANT: anteroventral, anteromedial, anterodorsal), ventral (ventral anterior, ventral lateral, ventral posterior), intralaminar (centromedian, centrolateral, parafascicular), medial (mediodorsal), and posterior (pulvinar, lateral posterior).

## Results

### Patient and Seizure Characteristics

Thirteen patients were included in the study: 10 from the sEEG group and three from the RC+S^TM^ group (Fig. 1). The median age of participants was 33 years (range: 20–65 years) (Supp. Table 1). There were no complications of thalamic electrode implantation in either group. Across both groups, 158 seizures were analyzed (Supp. Table 2).

**Figure 1.**
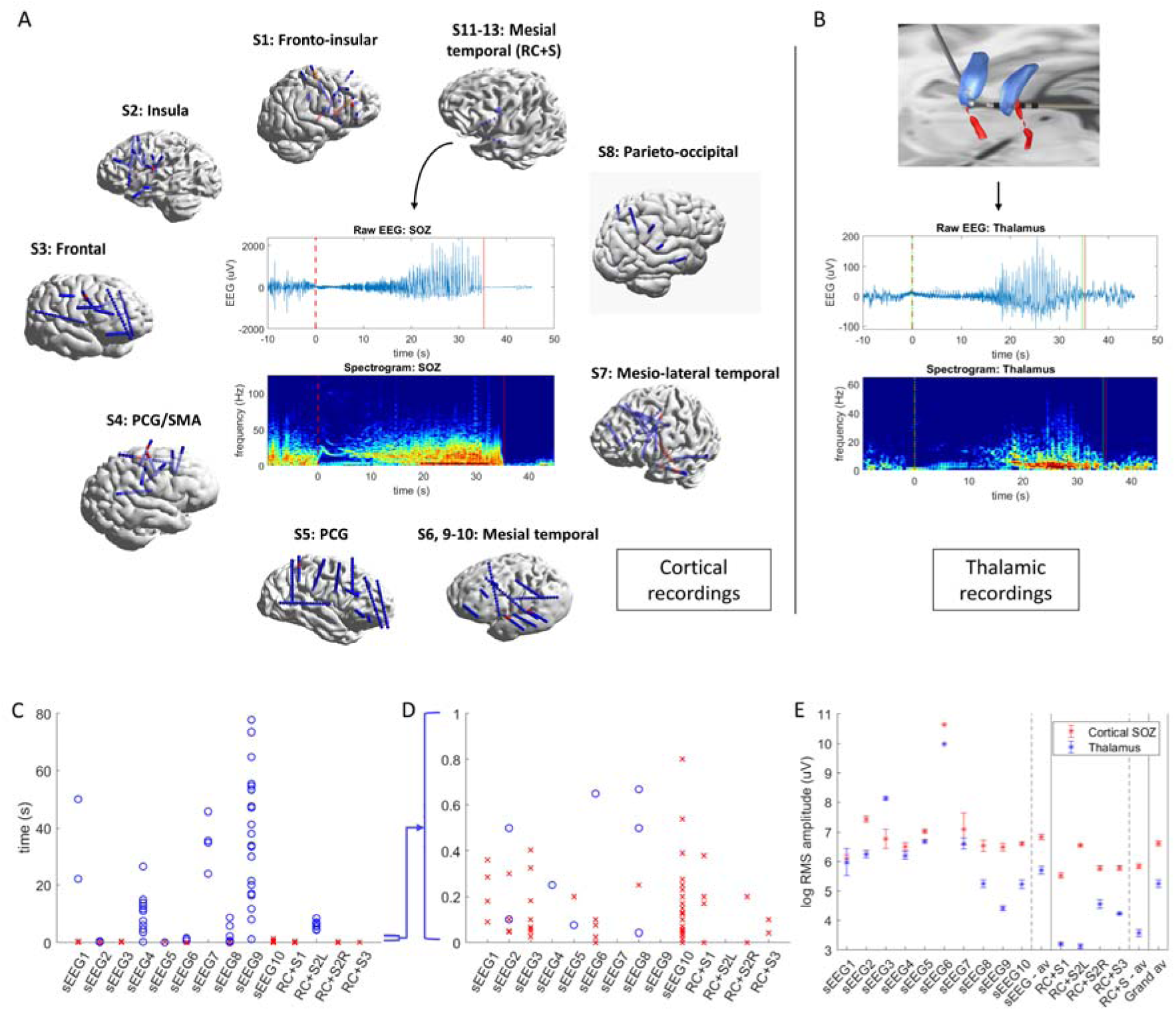
Simultaneous cortical and thalamic ictal onset recordings, corticothalamic onset delays, and ictal amplitude comparison. (**A**) 3D reconstructions of sEEG and chronic ambulatory device electrode implantations surrounding an example of the raw EEG recording from an exemplar cortical seizure onset zone (hippocampus), as well as time-frequency analysis of the cortical seizure onset zone discharge. **(B**) Top - 3D reconstruction of a thalamic electrode implantation targeting anterior thalamic nuclei bilaterally, parcellated and colored individually according to the Morel atlas. Blue - anteroventral (AV) nucleus; red – mammillothalamic tract. Bottom - example bipolar recording from the same anterior nucleus of the thalamus (AV) with time-frequency analysis of the thalamic ictal discharge. (**C** & **D**) Individual corticothalamic ictal onset delays are plotted for each seizure for each subject, sEEG1-10 and RC+S 1-3. All delays are shown in C, while panel D shows only the 0-1 second range. (**E**) Root mean square (RMS) amplitude of ictal discharge in cortical seizure onset zone (SOZ; in red) and thalamus (blue) are averaged and plotted for each participant, and averaged again across the two groups (sEEG and RC+S^TM^).

In the sEEG group, six patients had extra-temporal SOZ, with one case of multifocal extra-temporal onset. Two patients had temporal lobe SOZ, and two had multifocal temporal and extra-temporal SOZ. Thalamic sampling in this group included anterior (2/10), medial (4/10), intralaminar (6/10), ventral (6/10), and posterior (3/10) nuclei.

In the investigational device group, two patients had unilateral mesial temporal seizures recorded, while one patient exhibited bilateral mesial temporal seizure onsets. All seizures in this group were associated with anterior thalamic electrode recordings.

Across all patients, distinct thalamic ictal discharges were observed in at least one seizure. Of the 158 seizures analyzed, 140 (89%) demonstrated thalamic ictal discharges. Among these, 79 (56%) exhibited thalamocortical primary organization patterns, while the remainder were propagation patterns.

Corticothalamic delays ranged from near simultaneous (0-50 ms) to prolonged (60-90 seconds) (Fig. 1C, D; Supp. Fig 1). The median delay for primary onset patterns was 50 ms, while for propagation patterns the median was 12.3 s.

There were clear differences in amplitude of electrographic discharges in the thalamus as compared to the cortex (Fig. 1E; Supp. Fig 2). While there was significant individual variability, thalamic LFP seizure discharges were on average an order of magnitude lower in amplitude than cortical discharges.

### Thalamocortical Seizure Onset Patterns

Distinct thalamocortical patterns were observed, with seizure onsets classified as synchronous, early, or delayed (Fig. 2). Four characteristic thalamic onset patterns were identified, and listed along with how frequently they were observed:

1. Hypersynchronous Activity: Pre-ictal periodic or repetitive spiking, or sentinel spikes followed by seizure onset; 6/13 subjects.
2. Low-Voltage Fast Activity (LVFA): High-frequency LFP activity (>25 Hz); 7/13 subjects
3. Slow wave/Ictal baseline shift: A prominent LFP slow wave or baseline shift at onset; 3/13 subjects.
4. Suppression: Broad suppression of thalamic LFP background activity; 7/13 subjects.

**Figure 2.**
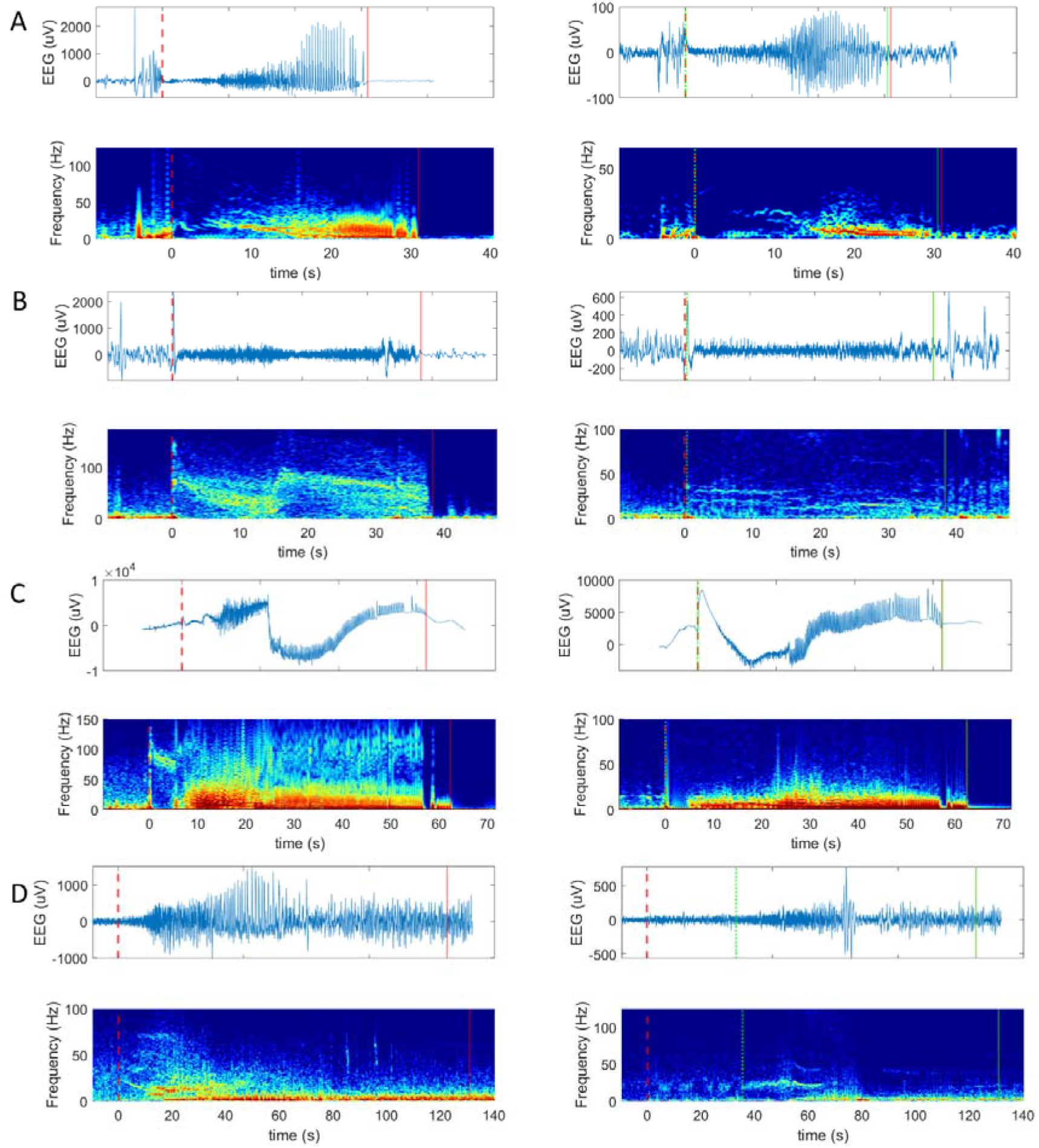
Corticothalamic propagation and seizure onset patterns. All panels: top row = raw EEG, bottom row is time-frequency plot (spectrogram); left column = cortical seizure onset zone, right column = thalamus. Dashed lines = seizure onset, solid lines =seizure offset; red = cortical discharge, green = thalamic discharge. (**A**): seizure onset in the mesial temporal region is essentially synchronous with the thalamus (anterior nucleus), with pre-ictal spiking followed by low-frequency suppression and low voltage fast activity seen in both mesial temporal and thalamic regions. (**B**): seizure onset in the posterior insula with synchronous thalamic discharge, consisting of hypersynchrony (large spike) followed by low frequency suppression and low-voltage fast activity. The same progression – pre-ictal spiking, ictal baseline shift, and LVFA is seen in the ventral thalamus (ventral lateral nucleus) albeit with slightly different spectro-temporal characteristics to the cortex. (**C**): seizure onset in the dorsolateral frontal lobe (middle frontal gyrus) is synchronous with thalamic onset (mediodorsal nucleus), the latter manifested as a large ictal baseline shift/slow wave with marked suppression of other frequencies, and eventually hypersynchrony in the form of repetitive spiking (no high pass filter in **C**, to allow for viewing of slow wave). (**D**): seizure onset occurs in the lateral temporal region, while thalamic activity (ventral posterior lateral nucleus) is delayed by over 30 seconds; representing a propagation pattern.

These patterns could occur in isolation or in or in combination. For example, hypersynchronous activity was frequently followed by LVFA, while baseline shifts often coincided with suppression of baseline LFP activity. Cortical and thalamic patterns were sometimes near identical but could also be distinct. For instance, LVFA in the cortex was often higher in frequency (50–150 Hz) and descended in frequency with time, compared to the corresponding thalamic LVFA (30–50 Hz), which often remained static with time. LVFA and suppression were the most common patterns (7/13 subjects each), and were seen together in 6/13 subjects. Anatomically, ictal baseline shifts were only seen in medial or ventral thalamus, while hypersynchronous activity was seen in all regions except medial thalamus (Supp. Table 3).

### Thalamocortical Propagation and Termination Patterns

Propagation patterns were characterized by delayed thalamic ictal discharges, often observed 10 – 60 seconds after cortical seizure onset (Fig. 1C; Fig. 2D; Supp. Fig. 1). These delayed discharges were associated with rhythmic ictal LFP oscillations (1–20 Hz) in thalamic contacts. In rare cases (4/13 patients; 5/158 seizures), thalamic discharges persisted beyond cortical seizure offset.

### Organisation of Thalamic Ictal Recruitment in Space and Time

Thalamic LFP activity during seizures varied across patients and seizure types, but was stereotyped for the same pattern of seizures within individual patients. Regional thalamic activation varied from no activation (Fig 3A), to focal-appearing activity (Fig 3B-C), to more diffuse changes (Fig. 3D). Broadband or low frequency suppression of thalamic LFP background rhythms was commonly present diffusely throughout the thalamus, often with superimposed more focal patterns (e.g. Fig 3C). Distinct spectral patterns could be observed to be spatially restricted (e.g. 1-2 electrode contacts), often correlated with a specific thalamic nucleus (e.g. ANT); and vary through time during a seizure.

**Figure 3.**
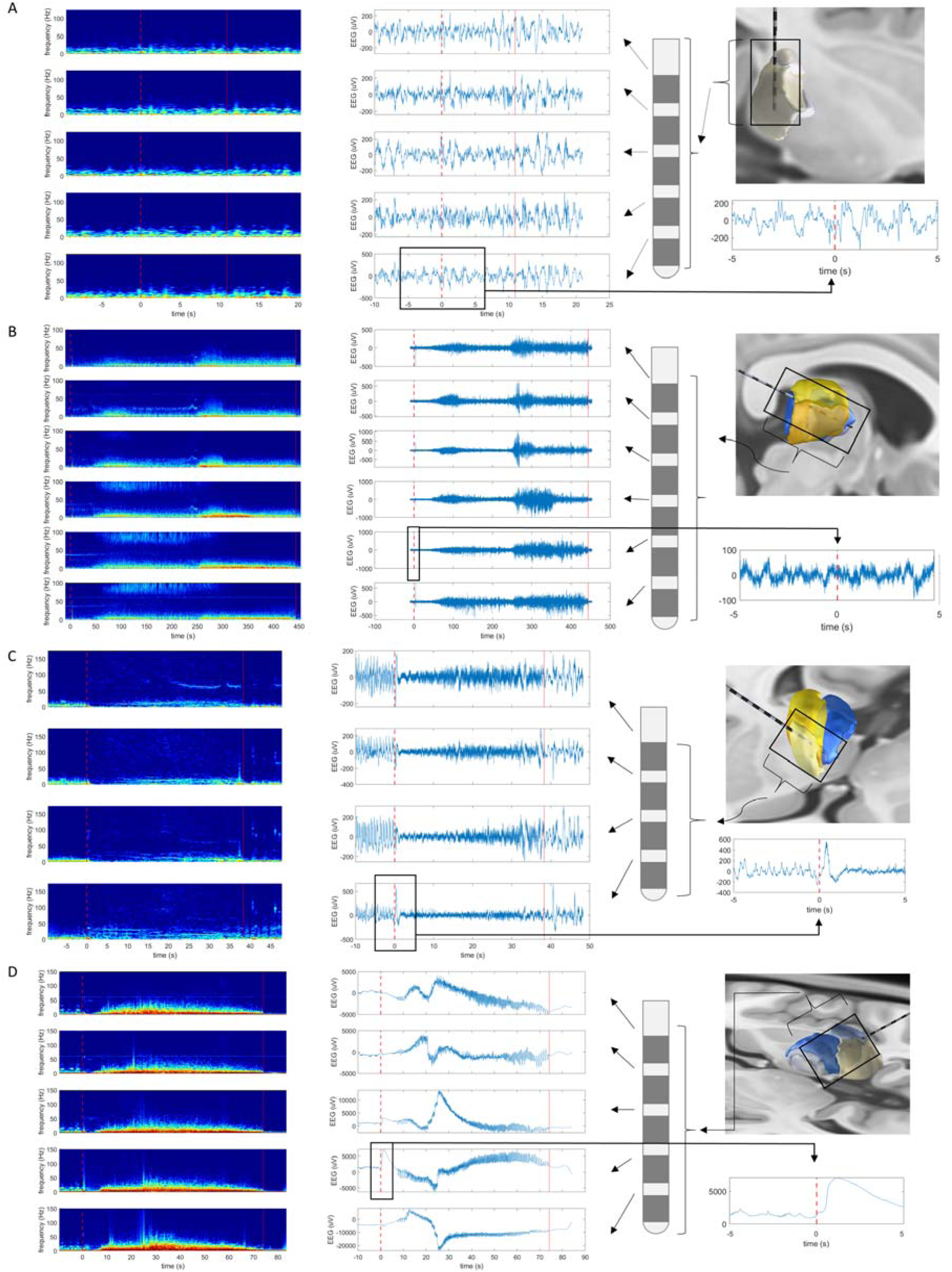
Thalamic ictal recruitment patterns. (**A**-**D**) all panels, left to right: time-frequency plot (left); raw EEG (middle); schematic of sEEG electrode with most distal contact at bottom of image, anatomical location of electrode, and zoomed in version of raw EEG (5 seconds before and after cortical seizure onset; (right). For each panel **A**-**D**: rows represent different bipolar recording channels in sampled thalamus. Dashed lines = seizure onset, solid lines = seizure offset; red = cortical discharge, green = thalamic discharge. Different thalamic recruitment patterns are shown: (**A**) no definite thalamic involvement; (**B**) long period of focal-appearing activity in a thalamic propagation pattern before more diffuse recruitment; (**C**) a diffuse suppression of background frequencies, with a more focal onset with the thalamus in the form of a large spike and low voltage fast activity restricted to a single thalamic channel, with evolution of recruitment of different thalamic regions over time; and (**D**) more ‘diffuse’ thalamic recruitment at seizure onset, with focality limited to a large ictal baseline shift/slow wave (no high pass filter in **D**, to allow for viewing of slow wave).

### Thalamocortical Seizure Circuits

The location of initial focal ictal activity within the thalamus varied according to the cortical SOZ. Characteristic thalamocortical seizure circuits were identified (Fig. 4), including:

- Frontal SOZ: Connected to medial, ventral, and intralaminar thalamus.
- Insular SOZ: Connected to ventral and intralaminar thalamus.
- Central/Peri-Rolandic SOZ: Connected to ventral and intralaminar thalamus.
- Parieto-Occipital SOZ: Connected to posterior thalamus.
- Lateral Temporal SOZ: Connected to medial and lateral thalamus.
- Mesial Temporal SOZ: Connected to anterior, medial, and ventral thalamus.

**Figure 4.**
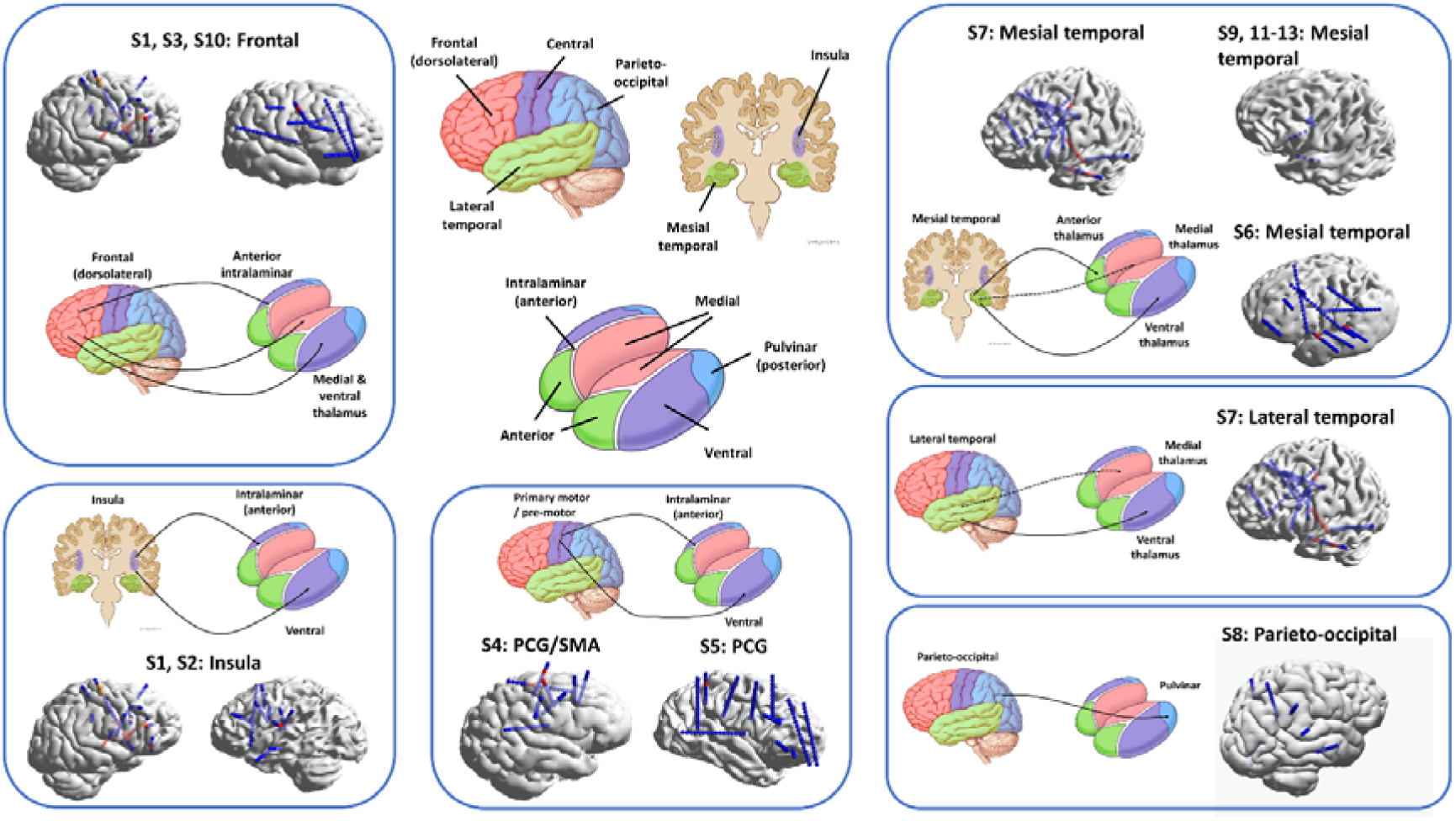
Corticothalamic seizure circuits. Circuits are grouped according to seizure onset zone and earliest thalamic recruitment. Clockwise from top left: frontal to intra-laminar, medial, & ventral thalamus; insula to intralaminar and ventral group; central/peri-rolandic to intralaminar/ventral groups; parieto-occipital to posterior (pulvinar) group; lateral temporal to medial/lateral groups; mesial temporal to anterior/medial/lateral groups. For each circuit, electrode positions (sEEG or RC+S^TM^) are shown in 3D reconstructed models, while circuit schematics depicting functional connectivity based on ictal EEG data are shown by arrows.

These circuits are largely consistent with known structural and functional thalamocortical networks, including the more diffuse projections associated with intralaminar nuclei.

## Discussion

Our findings highlight the central role of the thalamus in focal epilepsy, with ictal discharges observed in multiple thalamic nuclei across the majority of seizures. We observed thalamocortical networks at seizure onset characterized by thalamic activity that was simultaneous with cortical seizure onset and often paralleled this activity, suggesting a more intricate and complex thalamocortical dynamics underlying focal ictogenesis than is often assumed. More specifically, we identified primary organization patterns—characterized by distinctive synchronous (or near synchronous) thalamic discharges—and propagation patterns, in which thalamic activity followed cortical seizure onset by seconds to minutes. The involvement of diverse thalamic nuclei - including from the anterior, ventral, medial, posterior, and intralaminar groups - and multiple cortical connections of each nucleus, underscore the thalamus’s broad functional connectivity.

Distinct ictal onset patterns in the thalamus—hypersynchronous spiking, low-voltage fast activity (LVFA), ictal baseline shift, and suppression—were analogous to previously described cortical patterns,^22^ though with lower amplitudes and differing spectral characteristics. This suggests that the thalamic generators are closely coupled with cortical seizure activity while contributing unique dynamics to ictogenesis and propagation.

Similarly, we noted that thalamic ictal activity can be focal or diffuse and can evolve in space and time within intrinsic thalamocortical networks. This is analogous to these same well-described patterns in cortical seizure onset and propagation.

### Implications for Neurostimulation Therapies

Current DBS targets, such as the ANT and CMT, have been selected based on their connectivity to frontal and temporal networks and demonstrated efficacy in clinical trials.^2,4,6^ Our results suggest that other thalamic nuclei, including the pulvinar, medial, and other intralaminar thalamic nuclei, may also play key roles in focal epilepsy. Expanding DBS targets to include these regions, or combining stimulation at multiple thalamic nodes, could enhance therapeutic efficacy by suppressing onsets and propagation. Notably, we observed distinct thalamocortical circuits corresponding to different cortical SOZs. Tailoring stimulation targets based on individual thalamocortical networks may improve seizure control and reduce variability in patient outcomes. In addition, the classification of thalamic activity as focal or diffuse may confer prognostic information and inform the choice of focal thalamic stimulation (e.g. ANT-DBS) versus other forms of neurostimulation (e.g. VNS), similar to the way focality in SOZ influences decision-making in resective or ablative surgery.

Corticothalamic RNS approaches, which are being increasingly explored, require choosing electrodes for detection and stimulation. One strategy is to use a cortical lead for detection, given their presumed earlier role in focal seizure onset, while using a thalamic lead for stimulation. Our work suggests that appropriately placed thalamic leads could be used for both detection and stimulation, and the specific onset patterns we have identified could be used to choose and tune detectors for more accurate and timely seizure detection. This agrees and extends our other findings with RC+S^TM^ devices published by Gregg *et al.*^19^

### Implications for Epilepsy Diagnostics

The robust and stereotyped nature of thalamic ictal patterns raises the possibility that thalamic electrodes could serve as “sentinel” electrodes during sEEG evaluations, for example by lateralising the SOZ in rapidly propagating seizure networks, or identifying unsampled cortical regions contributing to seizure networks.

The differences in LFP amplitude and power spectral density observed in thalamus and cortex is related to the structural organization of the thalamus and biophysics properties of closed fields versus the open field of laminar structure in the cortex.^26^ The cortical columnar and laminar architecture generates higher amplitudes through summation, whereas the spherical/radial cellular nature of thalamic nuclei results in some cancellation of electric fields, and thus lower amplitudes and spectral power. The practical implication of this is that care must be taken in reading sEEG on standard sensitivities/gain, as low amplitude thalamic discharges may be missed. Similarly, removing high pass filters can allow for better visualization of ictal baseline shifts.

In addition to guiding patient selection and tailoring neurostimulation therapies through passive recordings, trial stimulation of thalamic targets during sEEG may help predict responses to chronic stimulation, further optimizing target selection for individual patients.^27^

Finally, there were no complications of thalamic sEEG implantation in this study, adding to the growing body of work supporting the safety of this procedure.^28^

### Pathophysiological Insights

This study reinforces the thalamus’s role as a hub in thalamocortical networks, dynamically linking cortical regions during ictogenesis and propagation. Mechanistically, the simultaneous and highly correlated nature of cortical and thalamic activity at focal seizure onset challenges the notion of the cortex as an independent focal seizure generator, at least in some cases. In generalised onset seizures, while undoubtedly seizures ‘start’ somewhere, the specific location probably varies, and is less important than existence of the underlying diffuse hyperexcitable network. In the same way, it may make less sense to talk about focal seizures ‘starting’ in the cortex, and instead preferable to consider hyperexcitable thalamocortical networks, which may range from very focal to more diffuse.

### Comparison with similar work

Our findings are comparable to those of Wu *et al.*^14^ who examined simultaneous cortical and thalamic sEEG findings in patients with presumed temporal lobe epilepsy. In this work, the first thalamic region activated was found to be variable, and to not always correspond to classical thalamocortical circuits. While we found similar variability, we sampled a broader range of nuclei and observed a stronger correlation of thalamic nucleus to cortical SOZ. Follow-up work also found an unexpectedly important role for PUL compared to ANT in temporal lobe seizures.^16^ Related analyses were undertaken in adults with temporal lobe epilepsy,^23^ and in a paediatric population with neocortical epilepsy,^29^ in which thalamic nuclei were sampled in addition to the seizure onset zone. The most common seizure onset pattern seen and analysed in these studies was low voltage fast activity, while we observed a range of patterns, analogous to those seen in the cortex.

### Limitations

A key limitation of this study is the sparse spatial sampling of thalamic nuclei, inherent to all intracranial evaluations. Electrode placements were guided by clinical hypotheses, leading to variable sampling across patients, and thalamic nuclei were not sampled systematically as some other studies have done. Additionally, anatomical localization of thalamic contacts is challenging due to the small size and close proximity of nuclei. Another limitation is the absence of surgical outcomes in most patients, precluding definitive confirmation of SOZ localization.

### Conclusion

This study highlights the critical role of the thalamus in the organization and propagation of focal seizures, providing insights into thalamocortical network dynamics in focal epilepsy. Our work contributes to the growing body of evidence supporting a network-based approach to epilepsy management and paves the way for novel diagnostic and therapeutic innovations.

## Abbreviations

ANT: anterior nucleus of the thalamus
CMT: centromedian nucleus of the thalamus
EBS: electrical brain stimulation
DBS: deep brain stimulation
DMT: dorsomedial nucleus of the thalamus
DRE: drug-resistant epilepsy
FDA: United States Food and Drug Administration
IRB: Institutional Review Board
LFP: local field potentials
LVFA: low voltage fast activity
PUL: pulvinar nucleus of the thalamus
RMS: root mean square
RNS: responsive neurostimulation
SOZ: seizure onset zone
sEEG: stereo-
EEG: stereotactic EEG
VNS: vagus nerve stimulation/stimulator.

## Data availability

The source data for the results presented in this manuscript are available from the authors on reasonable request.

## Acknowledgements

Artificial intelligence (a large language model based chatbot) was used in the preparation of the manuscript, to proof-read and summarise text in parts. It was not used for any part of reviewing the literature, analyzing or interpreting the data presented, or creating figures.

The authors would like to thank Certicon a.s. that provided EEG data viewer CyberPSG for research purposes.

## Supplementary material

The following supplementary files are available:

- Supplementary Material file: includes Supplementary Methods, Figures 1 & 2, and References
- Supplementary Tables 1, 2, 3

## References

1. Kwan P, Brodie MJ. Early identification of refractory epilepsy. New England Journal of Medicine. Published online 2000. doi:10.1056/NEJM200002033420503

2. Salanova V, Sperling MR, Gross RE, et al. The SANTÉ study at 10 years of follow-up: Effectiveness, safety, and sudden unexpected death in epilepsy. Epilepsia. 2021;62(6):1306–1317. doi:10.1111/EPI.16895

3. Nair DR, Laxer KD, Weber PB, et al. Nine-year prospective efficacy and safety of brain-responsive neurostimulation for focal epilepsy. Neurology. Published online 2020. doi:10.1212/WNL.0000000000010154

4. Simpson HD, Schulze-Bonhage A, Gregory |, et al. Practical considerations in epilepsy neurostimulation. Epilepsia. 2022;00:1–16. doi:10.1111/EPI.17329

5. Valentín A, García Navarrete E, Chelvarajah R, et al. Deep brain stimulation of the centromedian thalamic nucleus for the treatment of generalized and frontal epilepsies. Epilepsia. 2013;54(10):1823–1833. doi:10.1111/epi.12352

6. Dalic LJ, Warren AEL, Bulluss KJ, et al. DBS of thalamic centromedian nucleus for Lennox–Gastaut syndrome (ESTEL trial). Annals of Neurology. Published online 2021:4–4. doi:10.1002/ana.26280

7. Cukiert A, Cukiert CM, Burattini JA, Mariani PP. Seizure outcome during bilateral, continuous, thalamic centromedian nuclei deep brain stimulation in patients with generalized epilepsy: a prospective, open-label study. Seizure. 2020;81:304–309. doi:10.1016/j.seizure.2020.08.028

8. Burdette D, Mirro EA, Lawrence M, Patra SE. Brain-responsive corticothalamic stimulation in the pulvinar nucleus for the treatment of regional neocortical epilepsy: A case series. Epilepsia Open. 2021;6(3):611–617. doi:10.1002/EPI4.12524

9. Filipescu C, Lagarde S, Lambert I, et al. The effect of medial pulvinar stimulation on temporal lobe seizures. Epilepsia. 2019;60(4):e25–e30. doi:10.1111/epi.14677

10. Lundstrom BN, Osman GM, Starnes K, Gregg NM, Simpson HD. Emerging approaches in neurostimulation for epilepsy. Current Opinion in Neurology. 2023;36(2):69–76. doi:10.1097/WCO.0000000000001138

11. Herlopian A, Cash SS, Eskandar EM, Jennings T, Cole AJ. Responsive neurostimulation targeting anterior thalamic nucleus in generalized epilepsy. Annals of Clinical and Translational Neurology. 2019;6(10):2104–2109. doi:10.1002/acn3.50858

12. Tatum WO, Freund B, Middlebrooks EH, et al. CM-Pf deep brain stimulation in polyneuromodulation for epilepsy. Epileptic Disorders. n/a(n/a). doi:10.1002/epd2.20255

13. Nanda P, Sisterson N, Walton A, et al. Centromedian region thalamic responsive neurostimulation mitigates idiopathic generalized and multifocal epilepsy with focal to bilateral tonic–clonic seizures. Epilepsia. n/a(n/a). doi:10.1111/epi.18070

14. Wu TQ, Kaboodvand N, McGinn RJ, et al. Multisite thalamic recordings to characterize seizure propagation in the human brain. Brain. 2023;146(7):2792–2802. doi:10.1093/brain/awad121

15. Pizzo F, Roehri N, Giusiano B, et al. The Ictal Signature of Thalamus and Basal Ganglia in Focal Epilepsy: A SEEG Study. Neurology. 2021;96(2):e280–e293. doi:10.1212/WNL.0000000000011003

16. McGinn R, Von Stein EL, Datta A, et al. Ictal Involvement of the Pulvinar and the Anterior Nucleus of the Thalamus in Patients With Refractory Epilepsy. Neurology. 2024;103(11):e210039. doi:10.1212/WNL.0000000000210039

17. Romeo A, Roach ATI, Toth E, et al. Early ictal recruitment of midline thalamus in mesial temporal lobe epilepsy. Annals of Clinical and Translational Neurology. 2019;6(8):1552–1558. doi:10.1002/ACN3.50835

18. Pizarro D, Ilyas A, Chaitanya G, et al. Spectral organization of focal seizures within the thalamotemporal network. Annals of Clinical and Translational Neurology. 2019;6(9):1836–1848. doi:10.1002/ACN3.50880

19. Gregg NM, Marks VS, Sladky V, et al. Anterior nucleus of the thalamus seizure detection in ambulatory humans. Epilepsia. 2021;62(10):e158–e164. doi:10.1111/EPI.17047

20. Norden AD, Blumenfeld H. The role of subcortical structures in human epilepsy. Epilepsy & Behavior. 2002;3(3):219–231. doi:10.1016/S1525-5050(02)00029-X

21. Lüttjohann A, van Luijtelaar G. The role of thalamic nuclei in genetic generalized epilepsies. Epilepsy Research. 2022;182:106918. doi:10.1016/j.eplepsyres.2022.106918

22. Lagarde S, Buzori S, Trebuchon A, et al. The repertoire of seizure onset patterns in human focal epilepsies: Determinants and prognostic values. Epilepsia. 2019;60(1):85–95. doi:10.1111/EPI.14604

23. Ilyas A, Toth E, Chaitanya G, Riley K, Pati S. Ictal high-frequency activity in limbic thalamic nuclei varies with electrographic seizure-onset patterns in temporal lobe epilepsy. Clinical Neurophysiology. 2022;137:183–192. doi:10.1016/j.clinph.2022.01.134

24. Sladky V, Nejedly P, Mivalt F, et al. Distributed brain co-processor for tracking spikes, seizures and behaviour during electrical brain stimulation. Brain Communications. 2022;4(3). doi:10.1093/BRAINCOMMS/FCAC115

25. Horn A, Kühn AA. Lead-DBS: A toolbox for deep brain stimulation electrode localizations and visualizations. NeuroImage. 2015;107:127–135. doi:10.1016/j.neuroimage.2014.12.002

26. Einevoll GT, Kayser C, Logothetis NK, Panzeri S. Modelling and analysis of local field potentials for studying the function of cortical circuits. Nature Reviews Neuroscience 2013 14:11. 2013;14(11):770–785. doi:10.1038/nrn3599

27. Lundstrom BN, Gompel JV, Khadjevand F, Worrell G, Stead M. Chronic subthreshold cortical stimulation and stimulation-related EEG biomarkers for focal epilepsy. Brain Communications. 2019;1(1):fcz010. doi:10.1093/braincomms/fcz010

28. Gadot R, Korst G, Shofty B, Gavvala JR, Sheth SA. Thalamic stereoelectroencephalography in epilepsy surgery: a scoping literature review. Journal of Neurosurgery. 2022;1(aop):1–16. doi:10.3171/2022.1.JNS212613

29. Edmonds B, Miyakoshi M, Gianmaria Remore L, et al. Characteristics of ictal thalamic EEG in pediatric-onset neocortical focal epilepsy. Clinical Neurophysiology. 2023;154:116–125. doi:10.1016/j.clinph.2023.07.007

