## Supplementary Material for "Thalamocortical seizure onset patterns in drug resistant focal epilepsy"

**Supplementary Methods**

**Patient characteristics**

Patients were implanted with sEEG between October 2016 and March 2022; and with the investigational RC+S^TM^ device, between, November 2019 and April 2021. All patients were followed up from implantation until 25 November 2024.

The median age was 33 years (range: 20-65), the median follow-up time was 41 months (range 1-87 months), and the median seizure reduction at end of follow-up was 81% (range: 20-99%) (Supp. Table 1).

**sEEG Recordings**

Prolonged stereo-EEG (sEEG) recordings were acquired from the 10 patients as part of their routine inpatient clinical evaluation for drug resistant focal epilepsy. Lead electrode placements were determined by a multidisciplinary team of epileptologists, neuroradiologists, and neurosurgeons, targeting hypothesized seizure onset zones (SOZ). As described in the main text, in 10 patients, trajectories were extended into the thalamus to allow sparse sampling of thalamocortical networks involved in seizure generation and propagation, under IRB.

Stereo-EEG recordings were acquired using a Natus Xltek or Quantum system at a sampling rate of 256–500 Hz and filters 0.05 – 100 Hz.

**Investigational Device Recordings**

Three patients with mesial temporal lobe epilepsy were implanted with an investigational system (RC+S^TM^) with rechargeable battery and continuous LFP streaming and recording. Each patient had bilateral leads targeting amygdala/hippocampus and ANT. Signals were recorded at 250Hz sampling rate and filtered to (0.85-100 Hz band). Data were recorded over multiple months using a bipolar montage, and streamed to a cloud-based server via a tablet device. Seizures were automatically detected and confirmed through visual review by trained epileptologists as described previously.^1^

**Neurophysiologic data analysis**

Continuous local field potential (LFP) recordings were notch-filtered at 60 Hz, and additional high-pass filtering (>0.5 Hz) was applied (except where noted in relevant figures). Cortical and thalamic ictal discharges were classified based on time-domain and spectral features, including root mean square (RMS) amplitude and power-in-band analysis for delta (0–4 Hz), theta (4–8 Hz), alpha (8–13 Hz), beta (13–30 Hz), and gamma (30–100 Hz) frequency bands.

Thalamic seizure onset patterns were categorized into primary organization patterns (occurring within 0.5-1 s of cortical seizure onset, and analogous to those reported in cortex^2,3^) and propagation (non-specific) patterns. Descriptive statistics were calculated for seizure characteristics, thalamocortical delays, and power analysis. For each subject, cortical seizure onset zone is related to initial and maximal thalamic activation (Supp. Table 2). Circuit schematics were then constructed to associate thalamic nuclei with specific cortical SOZs.

**Neuroimaging**

Identification of specific thalamic nuclei was performed using Lead DBS and the Krauth/Morel atlas, adjusted for use with the Montreal Neurological Institute (MNI) 2009b asymmetric template space used in the Lead-DBS package.^4^

**Supplementary References**

1.	Sladky V, Nejedly P, Mivalt F, et al. Distributed brain co-processor for tracking spikes, seizures and behaviour during electrical brain stimulation. *Brain Communications*. 2022;4(3). doi:10.1093/BRAINCOMMS/FCAC115

2.	Lagarde S, Buzori S, Trebuchon A, et al. The repertoire of seizure onset patterns in human focal epilepsies: Determinants and prognostic values. *Epilepsia*. 2019;60(1):85-95. doi:10.1111/EPI.14604

3.	Pizzo F, Roehri N, Giusiano B, et al. The Ictal Signature of Thalamus and Basal Ganglia in Focal Epilepsy: A SEEG Study. *Neurology*. 2021;96(2):e280-e293. doi:10.1212/WNL.0000000000011003

4.	Horn A, Kühn AA. Lead-DBS: A toolbox for deep brain stimulation electrode localizations and visualizations. *NeuroImage*. 2015;107:127-135. doi:10.1016/j.neuroimage.2014.12.002

**Supplementary Figures**

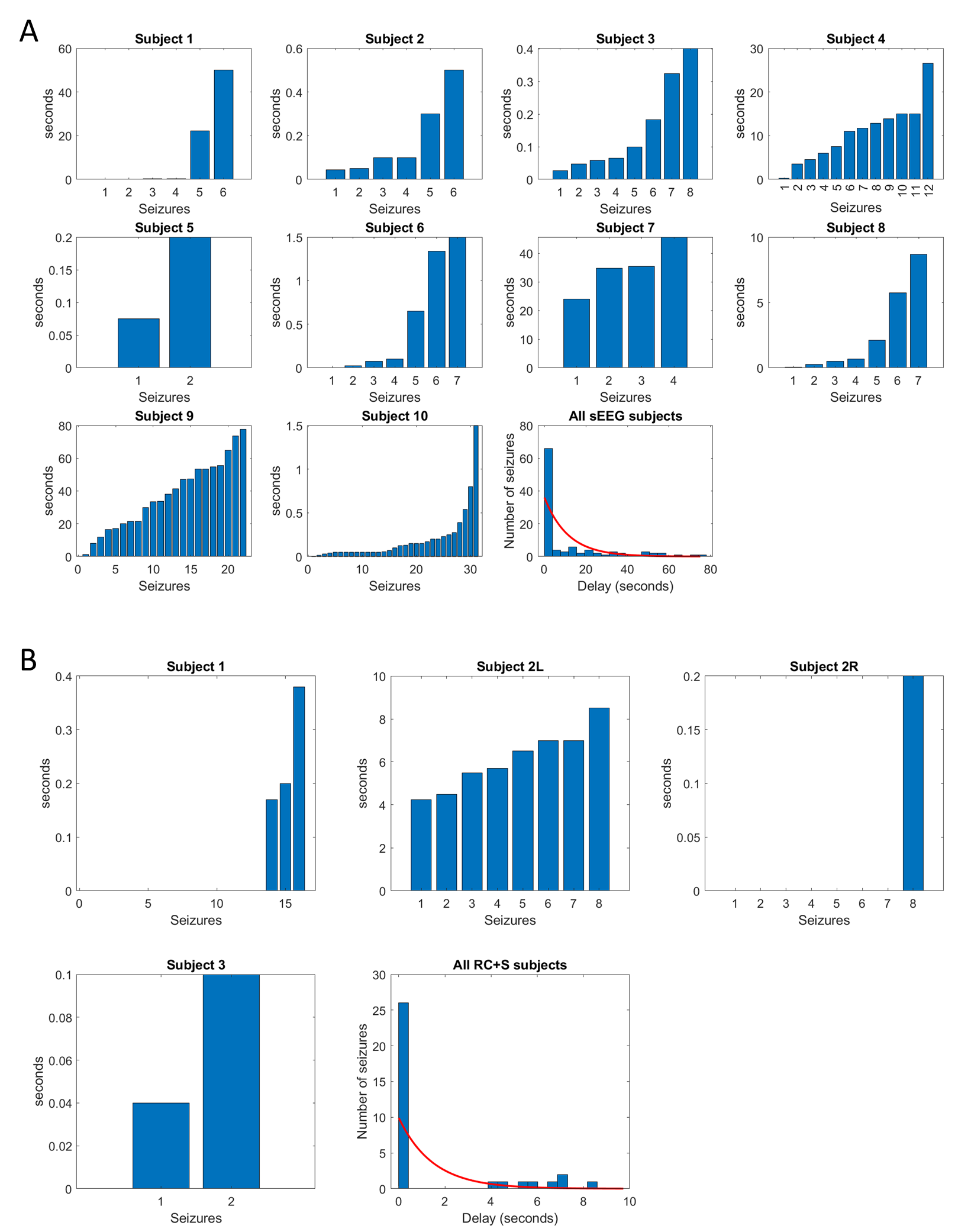

**Supplementary Figure 1. Cortico-thalamic ictal discharge delays.** (**A** & **B**) Delay distributions. Distributions of corticothalamic ictal onset delays are shown for each seizure in individual participants and averaged over all participants in **A** (sEEG) and **B** (RC+S^TM^). Red line: fitted (exponential) histogram of corticothalamic delays.

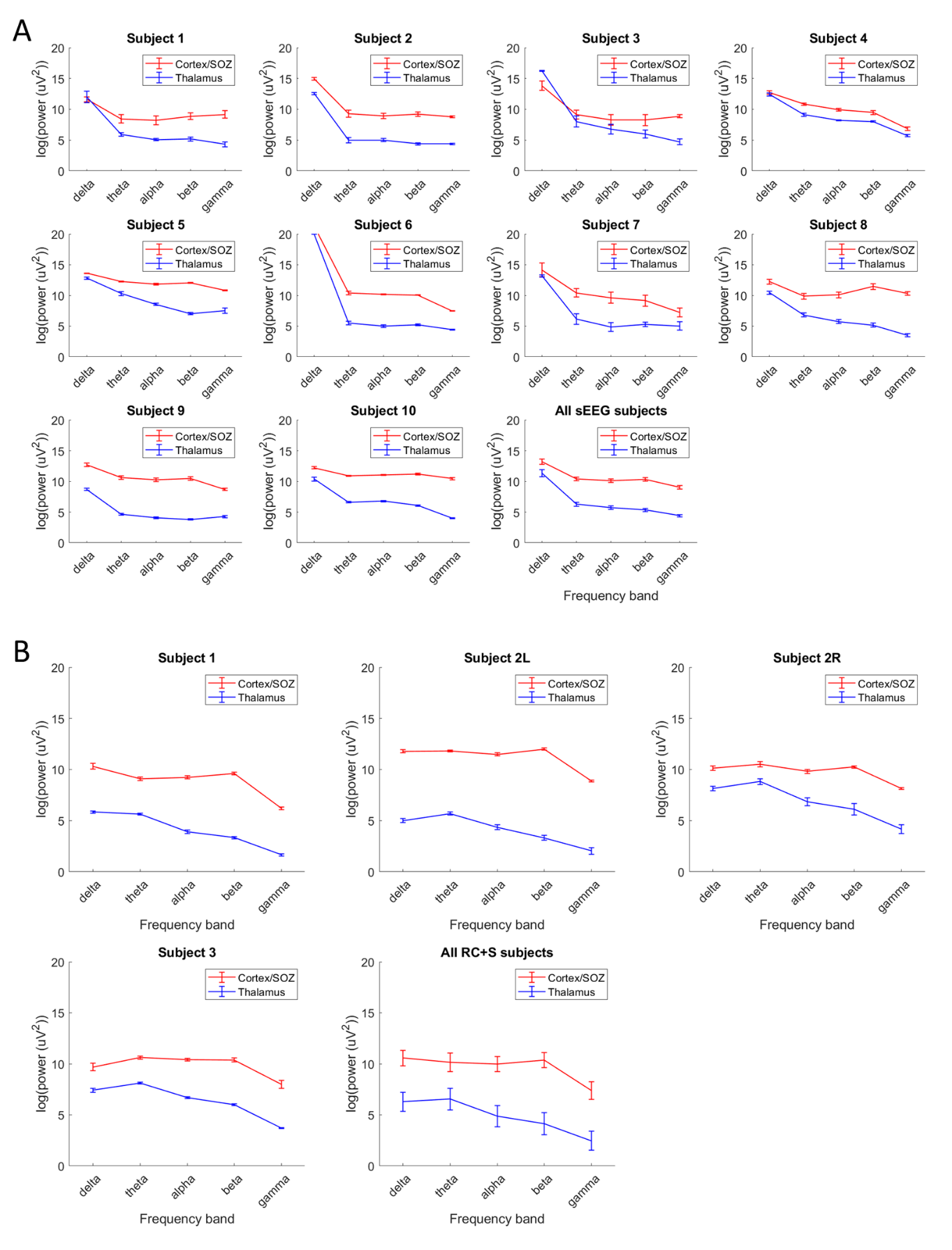

**Supplementary Figure 2. Cortico-thalamic ictal discharge voltage & power differences.** (**A** & **B**) Power in band. Power in band in the cortical seizure onset zone (red), and thalamus (blue) are shown for each participant averaged across their seizures, and averaged across participants in **A** (sEEG) and **B** (RC+S^TM^). Power in band is higher for all frequency bands on the group level and for most individuals, with the exception of lower (delta) frequencies in some participants.
