## Supplementary Table 1 for "Thalamocortical seizure onset patterns in drug resistant focal epilepsy"

**Supplementary Table 1. Clinical characteristics.**

| **Group / ID,**  **Sex (M/F)** | **Age (yrs.)**  **Epilepsy onset & implant** | **MRI**  **Normal (N) Lesion (L)** | **sEEG / DBS implantation complications** | **Temporal vs extratemporal** | **Intervention performed (after sEEG/passive recording)** | **Sz reduction at last F/U** | **Length of F/U (months)** |
| --- | --- | --- | --- | --- | --- | --- | --- |
| **sEEG** |  |  |  |  |  |  |  |
| *1 (M)* | 6-10; 21-25 | R hippocampal T2 signal change w/o atrophy; then S/P R ATL | None | Extratemporal | LITT, then CSS (insular x2, ANT, CMT) | 83% | 55 |
| *2 (M)* | 6-10; 26-30 | N | None | Extratemporal | None | N/A | 1 |
| *3 (M)* | 6-10; 16-20 | N | None | Extratemporal | Resection | 86% | 25 |
| *4 (F)* | 1-5; 31-35 | N | None | Extratemporal | Neuromodulation (CSS; SMA, precentral x2, postcentral) | 99% | 83 |
| *5 (F)* | 11-15; 26-30 | N | Lobar (parietal) hemorrhage (asymptomatic) | Extratemporal | Neuromodulation (CSS: ANT, 3 x peri-rolandic) | 20% | 87 |
| *6 (F)* | 56-60; 61-65 | Increased T2 signal / volume L hippocampus / amygdala | None | Temporal | Neuromodulation (RNS L hippocampus) | 93% | 57 |
| *7 (F)* | 26-30; 26-30 | N | None | Temporal & extratemporal | None | N/A | 9 |
| *8 (M)* | 6-10; 31-35 | L: left occipital encephalomalacia | None | Temporal & extratemporal | R HC LITT; L ANT + occipital RNS | 62.5% | 37 |
| *9 (F)* | 36-40; 36-40 | L: Left MTS | None | Temporal | L HC LITT; L HC FUS; L HC LITT | 82% | 38 |
| *10 (F)* | 6-10; 46-50 | N | None | Extratemporal | None | N/A | 17 |
| Median | 9.5; 29 |  |  |  |  |  | 37.5 |
| **RC+S^TM^** |  |  |  |  |  |  |  |
| *1 (F)* | 6-10; 56-60 | L: enlarged L amygdala / hippocampus | None | Temporal | ANT stimulation | 80% | 57 |
| *2 (F)* | 36-40; 41-45 | N*# | None | Temporal | ANT stimulation | 75% | 45 |
| *3 (F)* | 1-5; 31-35 | N* | None | Temporal | ANT stimulation | 57% | 41 |
| Median | 9; 45 |  |  |  |  |  | 45 |
| Grand median | 9; 33 |  |  |  |  |  | 41 |

*non-lesional MRI, but a history of anti-GAD antibody-associated autoimmunity is noted for these patients.

\# Vagus nerve stimulator (VNS) in situ prior to implant.

M = male; F = female; sEEG = stereo-EEG group; RC+S^TM^ = investigational device group; DBS = deep brain stimulation; F/U = follow-up; L = left; R = right; N = normal; MTS = mesial temporal sclerosis; S/P = status post; ATL = anterior temporal lobectomy; LITT = laser interstitial thermal therapy; CSS = chronic/continuous subthreshold stimulation therapy; ANT = anterior nucleus of the thalamus; CMT = centromedian nucleus of the thalamus; SMA = supplementary motor area; RNS = responsive neurostimulation; HC = hippocampus/hippocampal; FUS = (high intensity) focused ultrasound; NA = not applicable/not available.
