## Supplementary Table 2 for "Thalamocortical seizure onset patterns in drug resistant focal epilepsy"

**Supplementary Table 2. Seizure neurophysiology and semiology.**

| **Group** | **Subj.** | **Temporal vs extratemporal** | **Cortical seizure onset zone** | **Clinical manifestations** | **Thalamic discharge** | | **Thalamic nucleus (grouping)** | |
| --- | --- | --- | --- | --- | --- | --- | --- | --- |
|  |  |  |  |  | **Presence: n (%)** | **Type: n (%) primary** | **First change** | **Maximal activation** |
| *sEEG* | *1* |  |  |  |  |  |  | |
|  |  | Extratemporal | Anterior insula | None | 2/2 (100) | 0/2 (0) | Ventral anterior / lateral (VT) | Ventral anterior / lateral (VT) |
|  |  | Extratemporal | Posterior insula | Focal motor/FIAS | 2/2 (100) | 2/2 (100) | Centrolateral anterior (IT) | Ventral anterior / lateral (VT) |
|  |  | Extratemporal | Dorsolateral frontal (posterior orbital gyrus) | Focal motor/FIAS | 2/2 (100) | 2/2 (100) | Centrolateral anterior (IT) | Centrolateral anterior (IT) |
|  | *2* |  |  |  |  |  |  |  |
|  |  | Extratemporal | Posterior insula | Hypermotor (hyperkinetic) | 6/6 (100) | 4/6 (67) | Ventral lateral (VT) | Ventral lateral (VT) |
|  | *3* |  |  |  |  |  |  |  |
|  |  | Extratemporal | Frontal (dorsolateral) | Hypermotor | 6/6 (100) | 6/6 (100) | Mediodorsal (MT) | Pulvinar / lateral posterior (PT) |
|  |  | Extratemporal | Frontal (dorsolateral) | Hypermotor to bilateral tonic-clonic | 2/2 (100) | 2/2 (100) | Mediodorsal (MT) | Mediodorsal + centrolateral + pulvinar (MT/IT/PT) |
|  | *4* |  |  |  |  |  |  |  |
|  |  | Extratemporal | Frontal (precentral gyrus) | Subclinical/auras | 12/12 (100) | 0/12 (0) | Centrolateral anterior (IT) | Centrolateral anterior (IT) |
|  |  |  | Frontal (precentral gyrus) | Hemiclonic | 1/1 (100) | 0/1 (0) | Centrolateral anterior (IT) | Centrolateral anterior (IT) |
|  | *5* |  |  |  |  |  |  |  |
|  |  | Extratemporal | Frontal (precentral gyrus) | Focal motor | 2/2 (100) | 2/2 (100) | Ventral lateral (VT) | Ventral lateral (VT) |
|  | *6* |  |  |  |  |  |  |  |
|  |  | Temporal | Mesial temporal (amygdala) | Subclinical/minimally clinical | 3/3 (100) | 1/3 (33) | Ventral posterior lateral (VT) | Ventral posterior lateral (VT) |
|  |  | Temporal | Mesial temporal (amygdala) | Aphasia | 4/4 (100) | 3/4 (75) | Ventral posterior lateral (VT) | Ventral posterior lateral + centrolateral + mediodorsal (IT/MT/VT) |
|  | *7* |  |  |  |  |  |  |  |
|  |  | Temporal | Lateral temporal (ipsilateral) | Focal motor impaired awareness | 1/1 (100) | 0/1 (0) | Right ventral lateral / ventral posterior lateral (VT) | Right ventral lateral / ventral posterior lateral (VT) |
|  |  | Temporal | Lateral temporal (contralateral) | Focal motor impaired awareness | 2/2 (100) | 0/2 (0) | Right centrolateral (IT) | Right ventral lateral (VT) |
|  |  | Extratemporal | Posterior insula (contralateral) | Focal motor | 1/1 (100) | 0/1 (0) | Right centrolateral (IT) | Right ventral lateral (VT) |
|  | *8* | Extra-temporal | Parieto-occipital | Subclinical, visual auras | 6/10 (60) | 1/6 (17) | Pulvinar (PT) | Pulvinar (PT) |
|  |  | Extra-temporal | Parieto-occipital | FIAS | 1/1 (100) | 0/1 (0) | Pulvinar (PT) | Pulvinar (PT) |
|  |  | Temporal | Mesial temporal (contralateral) | FIAS | 1/1 (100) | 0/1 (0) | Pulvinar (PT) | Pulvinar (PT) |
|  | *9* | Temporal | Mesial temporal | Subclinical or FAS (sensory) | 22/36 (61) | 0/22 (0) | Anterior nucleus (AT) | Anterior nucleus (AT) |
|  | *10* | Extra-temporal | Dorsolateral frontal (middle frontal gyrus) | FIAS (behavioural/speech arrest, limb/trunk automatisms) | 30/30 (100) | 30/30 (100) | Ventral lateral (VT) | Ventral lateral (VT) |
|  | *Total* |  |  |  | 106/124 (85) | 53/106 (50) |  |  |
|  |  |  |  |  |  |  | **Prior sEEG** | **Thalamus chronic implant** |
| *RC+S^TM^* | *1* | Bitemporal  L >> R | L mesial temporal  R mesial temporal | FIAS (behavioural arrest, arousal from sleep)  None recorded/analysed | 16/16  NA | 16/16  NA | Yes (no thalamic sampling) | L & R ANT |
|  | *2* | Bitemporal  L ~= R | L mesial temporal  R mesial temporal | Subclinical  FIAS (with automatisms) | 8/8  8/8 | 0/8  8/8 | Yes (no thalamic sampling) | L & R ANT |
|  | *3* | Bitemporal  L >> R | L mesial temporal  R mesial temporal | FIAS; behavioural/speech arrest oral/manual automatisms  None recorded/analysed | 2/2  NA | 2/2  NA | No prior sEEG | L & R ANT |
|  | *Total* |  |  |  | 34/34 (100) | 26/34 (76) |  |  |
|  | *Grand total* |  |  |  | 140/158 (89) | 79/140 (56) |  |  |

M = male; F = female; sEEG = stereo-EEG group; RC+S^TM^ = investigational device group; DBS = deep brain stimulation; F/U = follow-up; L = left; R = right; AT = anterior thalamic group; VT = ventral thalamic group; IT = intralaminar thalamic group; MT = medial thalamic group; PT = posterior thalamic group; FAS = focal aware seizure; FIAS = focal impaired awareness seizure; ANT = anterior nucleus of the thalamus; HC = hippocampus/hippocampal; NA = not applicable/not available.
