## Supplementary Table 3 for "Thalamocortical seizure onset patterns in drug resistant focal epilepsy"

| **Subject** | **Thalamic location** | | **Hypersynchronous** | | | **LVFA** | **Baseline / DC shift** | **Suppression** |
| --- | --- | --- | --- | --- | --- | --- | --- | --- |
|  | **Group** | **Nucleus** | **Pre-ictal spiking** | **Sentinel spike** | **Repetitive spiking** |  |  |  |
| *1* | IT | CL | - | Y | - | Y | - | Y |
| *2* | VT | VL | - | - | - | Y | - | - |
| *3* | MT | MD | - | - | - | Y | Y | Y |
| *4* | IT | CL | - | - | - | - | - | - |
| *5* | VT | VL | Y | - | - | - | Y | Y |
| *6* | VT | VL/VPL | - | Y | - | Y | Y | Y |
| *7* | MT | CeM/MD | - | - | - | - | - | - |
| *8* | PT | PuM | - | - | - | - | - | - |
| *9* | AT | AV | - | - | - | - | - | - |
| *10* | VT | VL |  |  | Y |  |  |  |
| *11* | AT | AV | Y | - | - | Y | - | Y |
| *12* | AT | AV | Y | - | - | Y | - | Y |
| *13* | AT | AV | - | - | - | Y | - | Y |

**Supplementary Table 3. Thalamic sampling per subject and primary organisation pattern per thalamic region.**

AT = anterior thalamic group; VT = ventral thalamic group; IT = intralaminar thalamic group; MT = medial thalamic group; PT = posterior thalamic group; CL = centrolateral; VL = ventrolateral; MD = mediodorsal (dorsomedial); VPL = ventral posterior lateral; CeM = central medial; PuM = pulvinar, medial; AV = anteroventral; Y = yes.
